# Study protocol for the EYEdentify project: An examination of gaze behaviour in autistic adults using a virtual reality-based paradigm

**DOI:** 10.1101/2024.12.14.24319035

**Authors:** Alberte C. E. Jeppesen, Johannes Andresen, Rizwan Parvaiz, Lars Clemmensen, Jens Richardt Møllegaard Jepsen, Dan Witzner Hansen, Louise Birkedal Glenthøj

## Abstract

**Introduction:** Autism Spectrum Condition (ASC) is characterised by difficulties in social communication and interaction, which may pose significant challenges to daily functioning throughout life. While current diagnostic methods for ASC often rely on measures based on subjective reports, there is a growing need for objective, quantifiable measures to support current clinical assessment of ASC. Eye-tracking technology records eye and gaze movements in real time and provides a direct and objective method for assessing social attention. Integrating eye-tracking within virtual reality (VR) environments presents a novel approach for capturing gaze behaviour in dynamic, ecologically valid social scenarios. This study aims to investigate whether VR-based eye information can reveal group differences in gaze behaviour between autistic adults and neurotypical controls in simulated social interactions.

**Methods:** This case-control study will include 140 adults diagnosed with ASC and 50 neurotypical controls, matched by age and gender. Participants will engage in six VR-based social scenarios, which vary in social complexity and the presence of non-social distractors. Eye information will be measured using eye-tracking technology integrated into a head-mounted display. Gaze behaviour will be analysed through fixation-based metrics on parameters including number of fixations, mean fixation time, and dwell time, on predetermined Areas of Interest.

**Analysis:** Statistical analyses will assess between-group differences in gaze behaviour as well as correlations between gaze metrics and clinical measures of social functioning, social cognition and symptom severity.

**Discussion:** This study utilises VR-based eye-tracking to investigate novel paradigms for assessing gaze behaviour in ASC in immersive, interactive environments and aims to advance the current understanding of visual social attention in ASC. Positive outcomes from this study may support further research into VR- based eye-tracking to supplement existing clinical assessment methods.

## Introduction

*Autism Spectrum Disorder* (ASD) is a neurodevelopmental condition characterised by impairments in social communication and interaction, alongside restricted and repetitive behaviours and interests and deviant reactivity to sensory stimuli (1,2). In line with international recommendations, this paper will adopt the term *Autism Spectrum Condition* (ASC) and employ identity-first language (*autistic adults*), though it is acknowledged that preferences regarding terminology vary among individuals and communities (3–5).

Similarly, individuals without a developmental or current psychiatric diagnosis will be referred to as *neurotypical* individuals.

For autistic individuals, social-communicative impairments may present as challenges in social-emotional reciprocity, non-verbal communication, and difficulties forming relationships, often resulting in significant psychosocial distress and impacting daily functioning, occupational performance, and overall quality of life (6–8). The global prevalence of ASC is estimated to be between 1-2%, where adults constitute a growing proportion of this group (8,9). Being a neurodevelopmental condition, this increase in adult prevalence is unexpected, and several hypotheses have been proposed to account for the development, including revised and improved diagnostic criteria, increased awareness, and a growing recognition of compensatory strategies (*camouflaging*) that may have masked symptoms and resulted in children and adolescents going undiagnosed (8–12). As a result, “a lost generation” has formed, comprised of those individuals who remain undiagnosed or misdiagnosed into adulthood (13).

Diagnosis in adulthood presents unique challenges. As a neurodevelopmental condition, ASC requires that symptoms be present from childhood, meaning diagnosis is contingent on accurate retrospective reporting, often from family members, which may neither be accurate nor accessible (13). Additionally, many individuals who remain undiagnosed into adulthood do so as a result of acquiring compensatory strategies or camouflaging behaviours to mask their autistic traits in social settings, further complicating diagnostic processes (13,14). Current diagnostic methods are also reliant on subjective measures such as self-reports and clinical observations, which may be susceptible to bias, potentially leading to misdiagnosis (15).

Objective behavioural markers, such as gaze behaviour, may have the potential to support current clinical assessment methods for ASC.

Gaze behaviour provides a valuable behavioural measure in understanding social cognition. The development of eye-tracking technology has enabled the quantification of gaze patterns, and the study of how individuals allocate visual attention. In humans, gaze functions as a signal to modulate communication, evident in the attentional bias known as *visual social allocation*, where neurotypical populations tend to direct attention towards socially salient features, such as eyes and faces (16,17). Conversely, atypical gaze patterns may indicate aberrant social perception, reflected as impairments in social communication. As such, gaze behaviour may offer insights into assessing psychiatric conditions in which social communicative difficulties are present, such as ASC. The study of gaze behaviour in psychiatric populations has gained increasing attention in recent years, with studies examining its role in understanding various conditions, including ASC, social phobia, affective disorders and schizophrenia (18–22).

Atypical eye contact is frequently observed in autistic individuals and is recognised as a clinical feature in diagnostic classification systems (1,23). Studies suggest that autistic individuals process faces differently, directing less attention toward core facial features such as the eyes and nose (24), with a specific reduction in attention toward eyes regions (25,26), and, in some studies, increased attention towards mouth regions, compared to neurotypical controls (24,25). Meta-analyses corroborate the findings of reduced attention to eye regions in autistic individuals compared to neurotypical counterparts (26–28); however, findings concerning gaze patterns toward mouth regions remain inconsistent (26,27,29,30).

Beyond feature-specific differences in gaze patterns, autistic individuals also demonstrate a reduced preference for social regions (e.g. faces) compared to non-social regions (e.g. objects) within stimuli (26–29,31). Meta-analyses report that autistic individuals attend more to non-social regions of presented stimuli and direct less attention to socially relevant areas, such as whole-race regions, relative to neurotypical controls (26–29,31).

However, several moderating parameters within paradigms influence these observed differences. Dynamic stimuli, such as videos, tend to elicit greater differences between ASC and neurotypical groups when compared to static images (28,32,33). Additionally, social complexity within the stimuli, operationalised as the number of people present, impacts gaze behaviour (17,31,34). In neurotypical adults, increasing the social content of a stimulus leads to increased fixation time toward social regions, such as eyes and faces (17). Conversely, autistic individuals exhibit decreased attention to social regions, such as faces and eyes, and increased attention to non-social regions as social content increases (31,34).

Following this, much of the literature is limited due to the reliance on static or simplified stimuli, such as isolated images or videos, which fail to capture the interactive quality of real-world social situations (35). Passive viewing of stimuli also fails to account for the bi-directionality, or “dual function of gaze” in social situations, where in real-world interactions, gaze functions not only as a method of capturing information but also conveys communicative signals and intent (35). Gaze behaviour in a paradigm involving passive viewing may, therefore, not represent natural gaze behaviour, challenging the ecological validity of eye- tracking research that does not account for these factors.

Furthermore, a substantial part of the research investigating gaze behaviour in autistic individuals has been conducted in child and adolescent populations, highlighting the need for further studies examining gaze behaviour in adult ASC populations (28,29,31).

The use of virtual reality (VR) provides a promising avenue to counteract these limitations. VR technology enables the creation of highly immersive and complex stimuli resembling real-world situations while maintaining experimental control. Recent studies have demonstrated that participants exhibit stronger reactivity to the gaze of virtual avatars in VR settings compared to traditional laboratory settings, where the stimulus is presented on a monitor (36,37). Likewise, initial evidence suggests that combining a virtual environment eye-tracking paradigm with a machine learning model can differentiate autistic children from typically developing peers, with high (86%) accuracy and sensitivity (91%) based on differences in attuning to and extracting socially relevant information (38). This suggests that the use of VR-based stimuli in eye- tracking paradigms has the potential to increase ecological validity and contribute to the current understanding of visual social attention in autistic adults. The current study aims to expand on these preliminary findings by examining gaze behaviour in six computer-generated, immersive social scenarios, investigating whether VR-based eye-tracking can effectively identify group differences between autistic and neurotypical adults.

### Aim

This study aims to investigate the use of VR-integrated eye-tracking to assess gaze patterns in virtual social situations among autistic adults. More specifically, we intend to ascertain whether VR-based gaze patterns reveal group differences between autistic individuals and their neurotypical counterparts.

### Hypotheses

In line with previous findings (18,28,29,31–33), we hypothesise that:

1. Autistic participants will exhibit fewer fixations and shorter dwell time on eye regions of avatars compared to neurotypical participants across the experimental tasks.
2. Autistic participants will exhibit different gaze behaviours, such as fewer fixations and shorter fixation duration (dwell time) on social areas (head and body) of the avatars presented in the virtual reality (VR) scenarios compared to neurotypical participants. Additionally, we hypothesise that the number of fixations, mean fixation and dwell time will be greater on non-social areas (objects and surroundings) among autistic participants across the experimental tasks.
3. Increased social complexity across conditions, operationalised as a greater number of avatars present, will result in an increase in the number of fixations, mean fixation time, and dwell time on non-social areas among autistic participants compared to neurotypical controls.

## Methods

### Design

The study is a case-control study, including the baseline sample of 140 adults diagnosed with ASC enrolled in a randomised clinical trial (39) and a neurotypical control group of 50 adult individuals matched on age and sex. Participants will undergo a VR-based eye-tracking paradigm, wherein they will be presented with computer-generated stimuli within the *head-mounted display* (HMD). Gaze behaviour will be measured through eye-tracking technology integrated within the HMD. The study will employ a mixed-factorial design, featuring both within- and between-subject analyses.

### Setting

The study will be conducted by VIRTU Research Group at Mental Health Center Copenhagen, Copenhagen University Hospital, Denmark. Participants in the ASC group will be recruited from public and private outpatient diagnostic facilities in Denmark, specialised in the clinical assessment of ASC in adulthood, as well as from community-based social support services in the Capital Region of Denmark, provided they have a documented ASC diagnosis based on specialised clinical evaluation.

### Participants

#### Autism spectrum condition participants

A total of 140 autistic adults (ICD-10 F84; 84.1; 84.5; 84.8) will be included.

Inclusion criteria for the ASC group are as follows: (i) primary diagnosis of autism (ICD-10 F84; 84.1;84.5;84.8), (ii) age ≥ 18 years, (iii) a T-score on the SRS-A self-report of 60 or above, corresponding to 1 standard deviation above the general population mean, indicating clinically significant, mild to moderate difficulties with social interactions in daily life (40).

Exclusion criteria for the ASC group are as follows: (i) rejecting informed consent, (ii) a diagnosis of organic brain disease (ICD-10 F00 - F09), (iii) intellectual disability (IQ < 70), (iv) an insufficient command of spoken Danish or English, (v) an ADHD-diagnosis with medically untreated symptoms. ADHD symptoms are considered medically treated when participants demonstrate a satisfactory treatment response, as confirmed by their physician responsible for treatment. Additionally, participants must have no changes to dosage or type of medication within the preceding four weeks of study participation. This criterion aims to mitigate confounding influences and aligns with research indicating that medication-naïve adults with ADHD perform significantly worse than neurotypical controls on measures of visual attention, but that performance improves following six weeks of titrated methylphenidate treatment, including a two-week stable ‘end-point’ dosage (41). Given the high rate of psychiatric co-occurrence often present among individuals with ASC, co-occurring disorders will not constitute an exclusion criterion in the ASC group, apart from co-occurring ADHD with medically untreated symptoms (42,43).

#### Neurotypical participants

Fifty neurotypical participants will be recruited from the community, using advertisements at relevant institutions or through the websites www.forskningnu.dk and www.forsoegsperson.dk.

Inclusion criteria for the neurotypical group are as follows: (1) age ≥ 18 years, (2) ability to give informed consent.

Exclusion criteria for the neurotypical control group are as follows: (1) any current psychiatric diagnoses, including alcohol and drug dependence, (2) an insufficient command of spoken Danish or English.

Neurotypical participants will be matched to the ASC sample on age (+/- 2 years) and sex.

### Experimental paradigm

The experimental paradigm consists of six computer-generated social scenarios simulating a pedestrian street presented in virtual reality. Within each scenario, one avatar approaches the participant and initiates a 19- second interaction. The scenarios vary along two parameters: social complexity, operationalised as the number of avatars present in the background (0, 5, or 25), and the presence or absence of a non-social distractor (an ambulance in the background).

The virtual environment consists of a pedestrian street featuring elements such as buildings with large shopping windows, benches, and a large tree. The duration of each scenario is 90 seconds. When entering the scenario, participants are positioned centrally within the street, facing the same direction each time.

Participants can orient themselves while remaining stationary within the environment, i.e., they are unable to perform any locomotion. In each scenario, after 30 seconds, an interlocutor avatar emerges from a passage to the right of the participant’s initial orientation and approaches them. The interlocutor stops directly in front of the participant, adjusting to account for the participant’s physical orientation within the virtual space.

Once stopped, the avatar engages in a 19-second interaction involving an initial, neutral question, a brief pause allowing the participant to respond, and a neutral concluding remark (see Table 1 for an overview of the interactions). Following the interaction, the avatar exits the walking street down a passage located behind the participant’s original physical orientation. The avatars are distinct across scenarios, featuring three male and three female characters. All interacting avatars display consistent conversational body language and attempt to maintain eye contact. The scenario concludes after a total of 90 seconds.

**Table 1:**
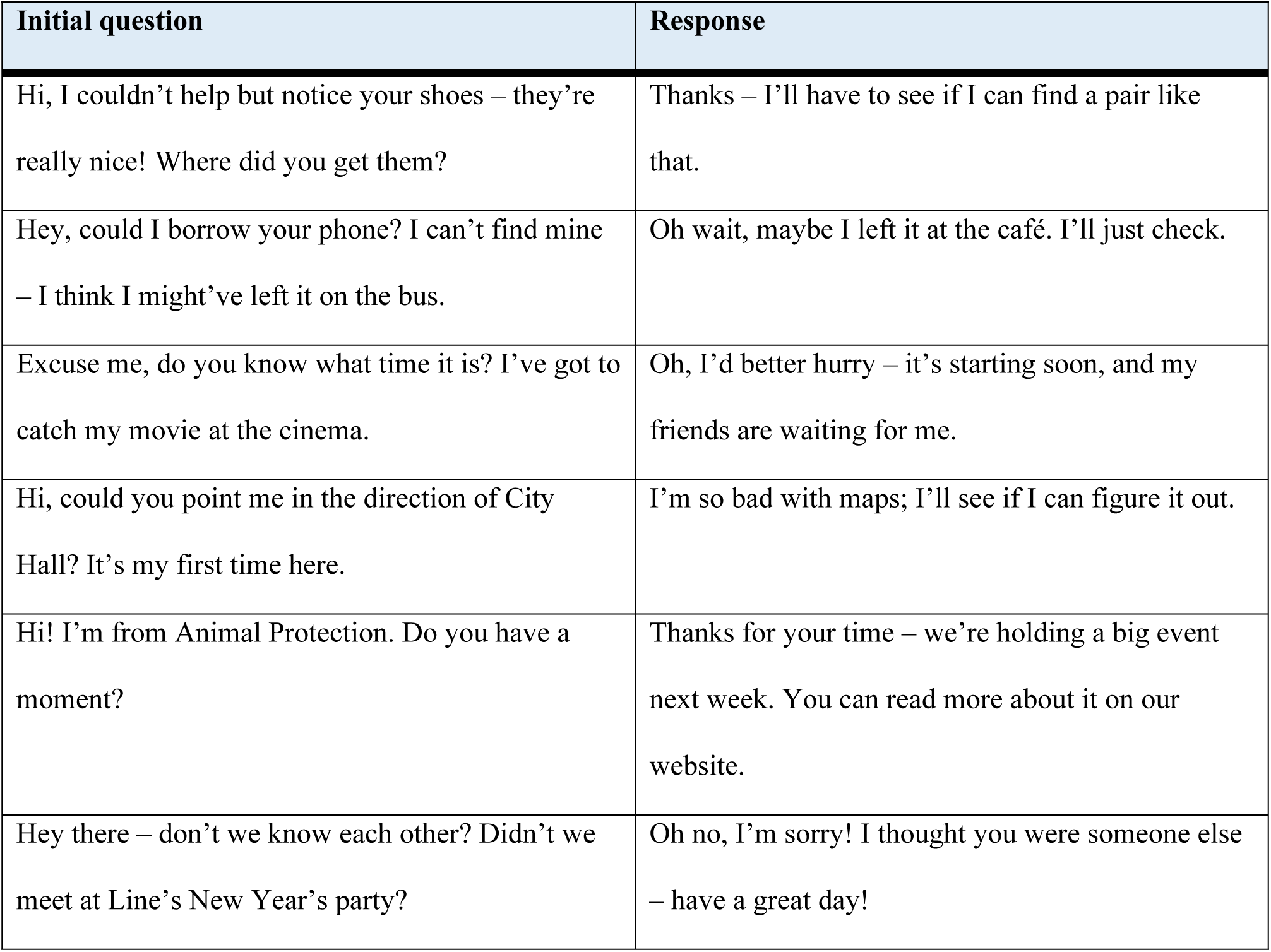
Overview of avatar interactions.

| Initial question | Response |
| --- | --- |
| Hi, I couldn't help but notice your shoes – they're really nice! Where did you get them? | Thanks – I'll have to see if I can find a pair like that. |
| Hey, could I borrow your phone? I can't find mine – I think I might've left it on the bus. | Oh wait, maybe I left it at the café. I'll just check. |
| Excuse me, do you know what time it is? I've got to catch my movie at the cinema. | Oh, I'd better hurry – it's starting soon, and my friends are waiting for me. |
| Hi, could you point me in the direction of City Hall? It's my first time here. | I'm so bad with maps; I'll see if I can figure it out. |
| Hi! I'm from Animal Protection. Do you have a moment? | Thanks for your time – we're holding a big event next week. You can read more about it on our website. |
| Hey there – don't we know each other? Didn't we meet at Line's New Year's party? | Oh no, I'm sorry! I thought you were someone else – have a great day! |

Across the six scenarios, two parameters are manipulated: social complexity and the presence of non-social distractors (see Table 2 for an overview of conditions for each scenario). Social complexity is operationalised as the number of avatars present in the background of each scenario, as this has been found to be a moderating factor when comparing gaze behaviour between ASC and neurotypical groups (31–33). Three conditions comprise this parameter: *1.* Low social complexity, with no avatars present within the background; *2.* Moderate social complexity, with five avatars walking in the background of the pedestrian street; *3.* high social complexity, with 25 avatars walking in the background on the pedestrian street. The second parameter, non-social distractors, is manipulated through the presence of an ambulance in half of the scenarios, as previous findings suggest that autistic individuals show preferential attention toward non-social objects in a visual scene (44–46).

**Table 2:** Experimental conditions.

|  | Non-social distractor absent<br>(Absent conditions) | Non-social distractor present<br>(Present conditions) |
| --- | --- | --- |
| Low social complexity<br>(0 background avatars) | Scenario 1 | Scenario 4 |
| Moderate social complexity<br>(5 background avatars) | Scenario 2 | Scenario 5 |
| High social complexity<br>(25 background avatars) | Scenario 3 | Scenario 6 |

### Apparatus

Stimulus presentation and gaze data collection will be conducted using the HMD Varjo Aero. Stimulus is presented through a high-resolution display of 2880 x 2720 pixels per eye, with a 115-degree horizontal field of view, at a 90 Hz refresh rate. Eye movements are recorded from both eyes at a sampling rate of 200 Hz with sub-degree accuracy. The Varjo Aero utilises a 5-point calibration technique. The HMD is paired with a high-performance PC, through which stimulus is controlled. Experimental stimuli are presented using the *Social World* software developed by CleVR. *Social World* was originally developed as a therapeutic tool in the treatment of various mental disorders, (47) but has been further developed to integrate an eye-tracking system.

### Procedure

Participants from both the ASC and neurotypical groups will be invited to the laboratory, informed about the purpose of the study, and complete an informed consent form. Prior to the experiment, participants undergo clinical assessment by experienced psychologists to ensure eligibility criteria are fulfilled and to collect clinical, cognitive, and behavioural measures (see section Assessment battery) to be included in the statistical analyses.

Following the clinical assessment, participants will proceed to the experimental task and receive verbal instructions about the general nature of the experimental task. Participants will be instructed to respond to the questions posed by the interacting avatars to increase immersion and presence within the task. Between each scenario, participants’ gaze will be calibrated through the 5-point calibration within the HMD to improve the accuracy of data collection. Following the experiment, participants will be debriefed and thanked for their participation.

### Assessment battery

Preceding the experimental task, all participants will undergo an assessment comprised of the following clinical and cognitive measures:

#### Clinical

- Mini International Neuropsychiatric Interview (M.I.N.I), v. 5.0 will be used to assess general psychopathology in order to record any cooccurring psychiatric conditions within the ASC group and to exclude any current psychiatric diagnoses within the neurotypical group (48).
- Danish Weschler Adult Intelligence Scale (WAIS-IV), subtests Vocabulary and Matrix reasoning, will be used to estimate full-scale IQ to confirm eligibility, which are known to correlate with full- scale IQ (49,50).
- Social Responsiveness Scale 2 – Adult Version, Self-report (SRS-2A Self-report), will be used to measure degree of social impairment (40).
- Social Responsiveness Scale 2 – Adult Version, Informant (SRS-2A Informant) will be used to measure degree of impairment as reported by an informant close to the participant (40)
- Social Interaction Anxiety Scale (SIAS) (51) will be used to measure levels of social anxiety.
- Empathy Quotient (EQ) will be used to measure level of empathy (52).
- Camouflaging Autistic Traits Questionnaire (CAT-Q) will be used to measure social camouflaging behaviours (12).
- Sensory Reactivity in Autism Spectrum questionnaire (SR-AS) will be used to assess degree of sensory reactivity, including both hyperreactivity and hyporeactivity (53).
- Beck Depression Inventory (BDI) will be used to assess level of depressive symptoms (54).
- Repetitive Behaviour Questionnaire (RBQ-3) will be used to assess repetitive behaviours, including motor stereotypies, ritualistic behaviours and restricted interests (55).
- Adult ADHD Self-Report Scale (ASRS) will be used to assess the severity of ADHD symptomology, including impulsivity-hyperactivity and inattention. (56)

#### Social functioning

- High Risk Social Challenge Task (HiSoC) is a performance-based measure and will be used to assess social functioning (57).
- Social Skills Performance Assessment (SSPA) is a performance-based measure and will be used to assess social functioning, including social appropriateness, conversational interest/disinterest and negotiation skills (58).
- Temple University Community Participation (TUCP) measure will be used to assess frequency and quality of community engagement over the course of the previous 30 days (59).
- Personal and Social Performance Scale (PSP) is used to assess level of social functioning, assessing functioning in domains including socially useful activities, personal and social relationships, self-care and aggressive or disturbing behaviours (60).

#### Social cognition

- CANTAB Emotion Recognition Task (CANTAB ERT) will assess the ability to identify emotions in presented images of facial expressions (61).
- The Assessment of Social Inference Test, Short form, part 2 social inference, minimal (TASIT-S) will assess theory of mind and complex emotion comprehension (62).
- The Hinting Task will assess theory of mind by measuring the ability to grasp indirect or implied meanings in social communication (63).

### Outcome measures

Outcome measures are gaze behaviour analysed through fixation-based metrics including number of fixations, mean fixation time in milliseconds, and dwell time in milliseconds on predetermined Areas of Interest (AOI). Areas of interest are predefined and categorised as social and non-social stimuli. Within social stimuli, AOIs are further categorised to include head, eye, mouth, forehead, and body regions of avatars. AOIs within non-social stimuli will consist of shopping windows, the ambulance when present, and the remaining background. Exploratory outcome measures are associations between eye movement data and clinical, social functioning, and social cognitive measures.

### Sample size calculation

Sample size was determined a priori based on one of the primary hypotheses of detecting a between-group difference in total fixation duration (dwell time) on the eye regions. The minimal detectable effect size as set at Hedges’ g = 0.47, as established by Frazier et al.’s 2017 meta-analysis (28). An allocation ratio of 0.35 was selected due to clinical availability to a cohort of participants with ASC (39) and anticipated greater heterogeneity in gaze behaviour within the ASC group (64).

Assuming a two-sided hypothesis with a significance level of 0.05 and statistical power of 80%, the sample size calculation indicated 139 and 48 NT participants. To account for potential attrition, we will recruit 140 ASC and 50 NT participants.

### Statistical analyses

Data will be analysed using Statistical Package for the Social Sciences (SPSS) and R software for statistical computing. Data analysis will be conducted using mixed design ANCOVA and MANCOVAs combining between-subjects and within-subjects analysis. Initially, a one-way ANOVA will be performed to examine group differences (ASC vs neurotypical) on demographic, clinical, psychosocial, and social cognitive measures. Means, standard deviations and F-scores will be reported.

To examine gaze behaviour across different regions of interest (social vs non-social areas and regions within these categories), MANCOVAs will be conducted with group (ASC vs neurotypical) as the between-subjects factor and AOIs as within-subjects factors. The potential influence of social complexity will be assessed using a MANCOVA with a group (ASC vs neurotypical) as a between-subject factor and social complexity and AOIs as within-subject factors. ASRS score will be included as a covariate to control for potential confounding influences of ADHD symptomology on visual attention.

Exploratory analyses using linear regression will evaluate potential correlations between eye-movement data and clinical, functional, and social cognitive measures.

All statistical tests for significance will be two-tailed and with a significance level alpha set at p<0.05. Any missing data will be handled using multiple imputation. In the event of non-normally distributed data, appropriate adjustments will be made, such as using corresponding non-parametric tests.

### Data management, safety consideration and ethics approval

The study will be conducted in accordance with the Helsinki Declaration and following the guidelines of the Danish Committee on Health Research Ethics. In compliance with the Data Protection Act and the Data Protection Regulation, patient information will only be collected with the individual’s consent. All participants will be provided with written and oral information. Written, informed consent is required for all the participants. Participants may at any time retract their consent without consequences. Electronic data from psychometric assessments will be stored pseudo-anonymized in the REDcap database, an online electronic data capture tool, meeting the requirements of the Danish Data Protection Agency. Eye-movement data will be stored pseudonymised in logged folders on network drives under the control of the Capital Region of Denmark, CIMT. Physical media, including consent forms, will be stored locally and securely.

Virtual reality is generally well tolerated, with minimal side effects or adverse events (65). Side effects may include experiences of cybersickness following time in VR (66). Side effects and adverse events will be monitored and recorded throughout the study. The study is registered at the Danish Data Protection Agency (p-2023-14488) and has been approved by the National Committee on Health Research Ethics in the Capital Region of Denmark (H-23055504).

### Status and timeline

Recruitment commenced in May 2024, with the first participant on May 2nd, 2024, and is expected to be completed in January 2026.

## Discussion

The proposed study aims to examine gaze behaviour differences between autistic adults and neurotypical controls using a VR-based eye-tracking paradigm. By investigating gaze behaviour within immersive, socially complex scenarios, this study aims to identify behavioural markers of ASC, which may hold clinical significance. Given that gaze abnormalities constitute a core feature of the clinical presentation of ASC (1), and are associated with difficulties in social communication and interaction, the findings from this study could expand on the current understanding of underlying social-cognitive deficits in ASC. The results may serve as the foundation for future studies investigating VR-based eye-tracking as an objective, non-invasive tool for assessing visual social attention in ASC.

A significant clinical implication of this research is the potential for developing objective assessment tools to aid in the clinical assessment of ASC. Eye-tracking technology, when integrated with VR, may offer an enhanced, ecologically valid method for evaluating gaze behaviour in real-world social contexts, thereby supporting current clinical assessment methods (20,37,38). Although there are existing tools to evaluate social deficits in ASC, such as standardised behavioural assessments, these often rely on subjective clinician observations or self-report measures. VR-based eye-tracking offers the opportunity to observe gaze behaviour in a controlled, ecologically valid environment, providing data that may supplement existing diagnostic procedures.

Aberrant gaze patterns have been observed in a range of psychiatric conditions, such as affective disorders and schizophrenia (21,22,67). As the current study focuses exclusively on an ASC population, future research may examine whether specific gaze patterns are unique to ASC or may be shared across clinical populations.

While VR technology is increasingly employed in therapeutic interventions (47,66,68,69), its use as a diagnostic or assessment tool remains relatively underexplored (70). By measuring gaze behaviour in VR- based paradigms, this study may contribute to the potential utility of VR as an instrument when assessing psychopathology. Findings may also have therapeutic implications if distinct gaze patterns are identified within the ASC group, where gaze avoidance may become focal points for therapeutic interventions.

## Data Availability

No datasets were generated or analysed during the current study. All relevant data from this study will be made available upon study completion.

## Acknowledgements

The authors extend their sincere gratitude to the psychiatric clinics and community-based social support services that will assist with the recruitment of participants for the study.

